# Performance of o1 pro and GPT-4 in self-assessment questions for nephrology board renewal

**DOI:** 10.1101/2025.01.14.25320525

**Authors:** Ryunosuke Noda, Chiaki Yuasa, Fumiya Kitano, Daisuke Ichikawa, Yugo Shibagaki

**Author notes:** Corresponding author: Ryunosuke Noda, The address of all authors: 2-16-1 Sugao, Miyamae-Ku, Kawasaki, Kanagawa 216-8511, Japan.

## Abstract

**Background:** Large language models (LLMs) are increasingly evaluated in medical education and clinical decision support, but their performance in highly specialized fields, such as nephrology, is not well established. We compared two advanced LLMs, GPT-4 and the newly released o1 pro, on comprehensive nephrology board renewal examinations.

**Methods:** We administered 209 Japanese Self-Assessment Questions for Nephrology Board Renewal from 2014–2023 to o1 pro and GPT-4 using ChatGPT pro. Each question, including images, was presented in separate chat sessions to prevent contextual carryover. Questions were classified by taxonomy (recall/interpretation/problem-solving), question type (general/clinical), image inclusion, and nephrology subspecialty. We calculated the proportion of correct answers and compared performances using chi-square or Fisher’s exact tests.

**Results:** Overall, o1 pro scored 81.3% (170/209), significantly higher than GPT-4’s 51.2% (107/209; p<0.001). o1 pro exceeded the 60% passing criterion every year, while GPT-4 achieved this in only two out of the ten years. Across taxonomy levels, question types, and the presence of images, o1 pro consistently outperformed GPT-4 (p<0.05 for multiple comparisons). Performance differences were also significant in several nephrology subspecialties, such as chronic kidney disease, confirming o1 pro’s broad superiority.

**Conclusion:** o1 pro substantially outperformed GPT-4 in a comprehensive nephrology board renewal examination, demonstrating advanced reasoning and integration of specialized knowledge. These findings highlight the potential of next-generation LLMs as valuable tools in specialty medical education and possibly clinical support in nephrology, warranting further and careful validation.

## INTRODUCTION

Recent advances in artificial intelligence, particularly in large language models (LLMs), have dramatically improved their capabilities. By learning from vast amounts of data, LLMs have achieved an unprecedented level of language comprehension, including the ability to organize and summarize extensive information [1]. Their accessibility— requiring no specialized programming skills and simulating the experience of interacting with an expert consultant—has made them increasingly appealing to clinicians. This progress has expanded their potential use across healthcare, from medical education and clinical practice to research support [2–6]. In nephrology, for example, LLMs are being investigated for dietary counseling in kidney disease [7], tailoring hemodialysis prescriptions [8], enhancing kidney transplant care [9], and supporting nephrology-specific literature retrieval [10].

Among LLMs, the Generative Pretrained Transformer (GPT) series from OpenAI has garnered particular attention [11]. These models demonstrate strong adaptability to English and multilingual environments, excelling in various medical assessments— including national medical licensing examinations in Japan and the United States, and specialty exams [12–17]. Notably, GPT-4, a flagship model in this series [18], reportedly achieved passing scores on the Polish nephrology board examinations [19], suggesting that LLMs might perform well even in highly specialized medical fields such as nephrology.

The field of LLMs continues to evolve. In December 2024, OpenAI released the o1 series—models developed through advanced reinforcement learning that exhibited enhanced reasoning abilities and fewer instances of hallucinations (factually ungrounded responses) [20]. In medical fields, o1 achieved over 90% accuracy on the benchmark consisting of Japanese national medical licensing examinations [21]. Its enhanced version, o1 pro, reportedly performed even better in domains requiring rigorous scientific reasoning in mathematics, science, and coding [22]. The emergence of o1 pro marks the next generation of LLMs, potentially surpassing GPT-4, thereby heightening expectations for specialized applications.

Despite these advances, it remains unclear whether LLMs can handle the rigor, complexity, and contextual demands of nephrology—a field requiring integrated clinical reasoning, specialized knowledge, and the interpretation of complex data, including imaging and pathology. The Self-Assessment Questions for Nephrology Board Renewal (SAQ-NBR), provided by the Japanese Society of Nephrology, offer a comprehensive set of Japanese-language multiple-choice nephrology questions [23]. These encompass fundamental concepts and complex clinical scenarios, including image-based challenges (kidney biopsies, imaging findings), and cover a wide range of nephrology subspecialties. Passing the SAQ-NBR requires integrative knowledge and clinical reasoning, making it a strict benchmark for advanced nephrology competence.

This study compared the performance of GPT-4 and the newly released o1 pro on the SAQ-NBR, clarifying whether LLMs could meet the cognitive and domain-specific challenges posed by complex nephrology content. Through this comparison, we aimed to provide insights into their potential applications in nephrology education and clinical support.

## MATERIALS AND METHODS

### The Self-Assessment Questions for Nephrology Board Renewal

The Self-Assessment Questions for Nephrology Board Renewal (SAQ-NBR) are a series of multiple-choice questions administered annually by the Japanese Society of Nephrology [23,24]. These questions, presented in Japanese, serve as a reference tool for nephrologists seeking board renewal. The passing criterion is defined as achieving ≥60% correct answers. Each year’s examination comprises a range of clinical and basic science questions that collectively span the breadth of nephrology. A subset of the questions includes images—such as renal biopsy specimens and radiological images—to assess interpretive and diagnostic skills. In this study, we included a total of 209 SAQ-NBR items from examinations between 2014 and 2023, excluding a single question that had been officially withdrawn as invalid by the Society.

### Classification by Taxonomy, Question Type, Image Inclusion, and Subspecialty

Following a previous report [16], we classified each question into four categories: Taxonomy: Based on the question creation manual for the Japanese National Medical Examination created by the Japan Medical Association [25], questions were categorized into three cognitive levels: recall, interpretation, and problem-solving. This taxonomy reflects the escalating depth of cognitive processing required to arrive at the correct answer.

Question Type: Questions were divided into general (focusing on fundamental medical or nephrological knowledge) and clinical (requiring clinical decision-making or patient management strategies).

Image Inclusion: Questions were designated as either image-based (image questions), incorporating visual data such as histopathological or radiological findings, or text-only (non-image questions).

Subspecialty: Drawing on the classification scheme of the Japanese Society of Nephrology’s case experience list [26], questions were assigned to one of several nephrology subspecialty areas: chronic kidney disease/end-stage kidney disease (CKD/ESKD), acute kidney injury (AKI), glomerular diseases, tubulointerstitial diseases, hypertension/vascular diseases, water/electrolyte/acid-base disorders, autosomal dominant polycystic kidney disease (ADPKD)/urology, or basic science.

### LLM models (o1 pro and GPT-4)

The o1 pro model, released in December 2024, represents the latest generation of LLMs, purportedly offering superior reasoning capabilities [20]. GPT-4, introduced in March 2023, has demonstrated high performance on various medical examinations and has been widely recognized for its medical reasoning prowess [3,18]. We accessed both o1 pro and GPT-4 through ChatGPT pro interface. All prompts were input in December 2024.

To prevent context learning, each question was presented in a new chat session. Exceptionally, when consecutive questions pertained to the same clinical case, the chat session was not refreshed, and the same session was used to input subsequent questions. For each prompt, we stated in Japanese: “We will now present a nephrology-related question. Please provide your answer and explanation.” The full text of the question was then provided. For image-based questions, the corresponding image was captured as a PNG file using the standard Windows screenshot tool and input simultaneously with the question text. The responses from both models were recorded, and their correctness was adjudicated based on the official answers provided by the Japanese Society of Nephrology.

### Statistical Analysis

For both o1 pro and GPT-4, the overall proportion of correct answers for ten years and the proportion of correct answers by year were calculated, and whether they met the pass criteria (≥60% correct) for each year was evaluated. Additionally, we calculated and compared correct answer proportions by taxonomy, question type, image inclusion, and nephrology subspecialty. Statistical analyses were performed using Python version 3.10.12. Differences in proportions were evaluated using chi-square or Fisher’s exact tests, as appropriate. A p-value <0.05 was considered statistically significant.

## RESULTS

### Overall and Annual Performance

Across the 209 SAQ-NBR questions between 2014 and 2023, we confirmed the absence of duplicate content. The distribution of the number of questions and their classifications by year is detailed in Supplemental Table S1. Overall, o1 pro achieved a proportion of correct answers of 81.3% (170/209), significantly surpassing GPT-4’s 51.2% (107/209; p<0.001, Table 1). When examined by year, o1 pro consistently maintained high accuracy (70–95%) and exceeded the ≥60% threshold in every examination year. In contrast, GPT-4 showed considerable variability (35–72%) and passed the 60% threshold in only two of the ten years. Notably, in the 2016, 2018, and 2020 examinations, o1 pro recorded accuracy of 95.0%, 95.0%, and 84.2%, respectively, significantly outperforming GPT-4’s 55.0%, 55.0%, and 31.6% for the corresponding years (p=0.011, 0.011, 0.003).

**Table 1:**
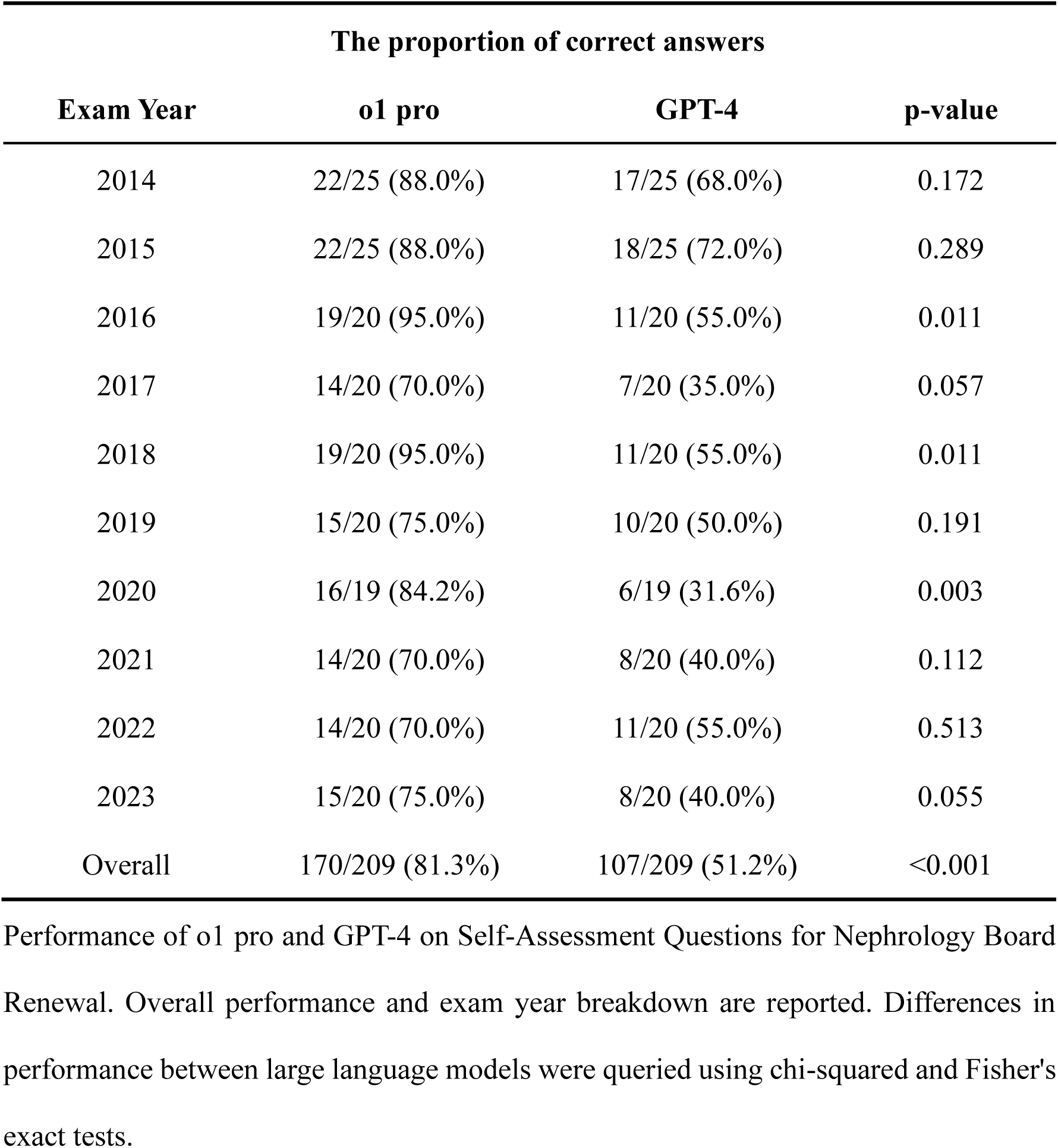
The proportion of correct answers of o1 pro and GPT-4 by exam year on the Self-Assessment Questions for Nephrology Board Renewal.

### Category-Specific Performance

By taxonomy, o1 pro significantly outperformed GPT-4 across all cognitive levels (Table 2). For recall-level questions, o1 pro achieved an 83.3% accuracy (90/108) compared to GPT-4’s 49.1% (53/108; p<0.001). For interpretation-level questions, the respective rates were 75.0% (42/56) versus 50.0% (28/56; p=0.011). At the most complex level, problem-solving questions, o1 pro maintained an advantage with 84.4% (38/45) versus 57.8% (26/45; p=0.011).

**Table 2:**
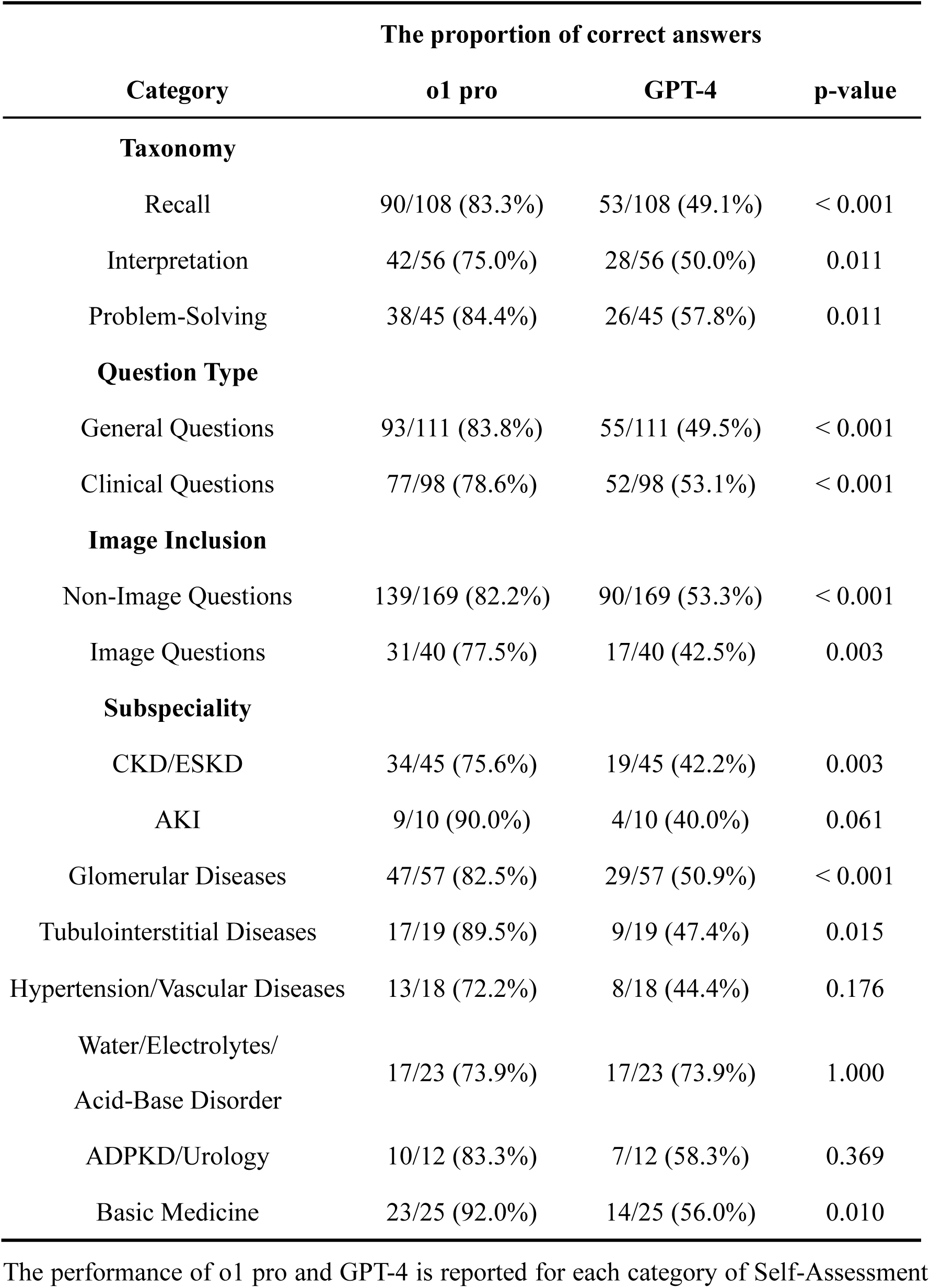
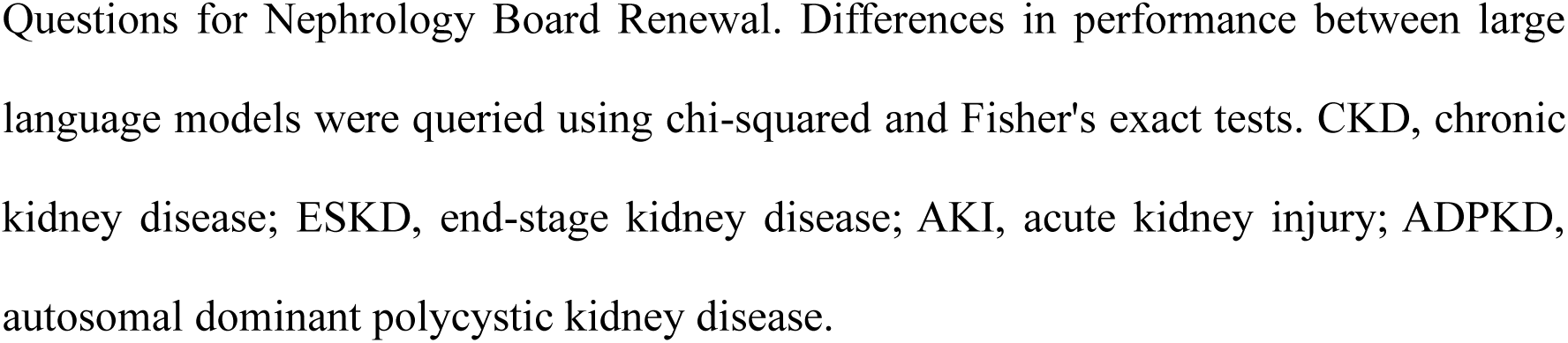
The proportion of correct answers of o1 pro and GPT-4 by four categories on the Self-Assessment Questions for Nephrology Board Renewal.

| Category | The proportion of correct answers |  |  |
| --- | --- | --- | --- |
|  | o1 pro | GPT-4 | p-value |
| **Taxonomy** |  |  |  |
| Recall | 90/108 (83.3%) | 53/108 (49.1%) | < 0.001 |
| Interpretation | 42/56 (75.0%) | 28/56 (50.0%) | 0.011 |
| Problem-Solving | 38/45 (84.4%) | 26/45 (57.8%) | 0.011 |
| **Question Type** |  |  |  |
| General Questions | 93/111 (83.8%) | 55/111 (49.5%) | < 0.001 |
| Clinical Questions | 77/98 (78.6%) | 52/98 (53.1%) | < 0.001 |
| **Image Inclusion** |  |  |  |
| Non-Image Questions | 139/169 (82.2%) | 90/169 (53.3%) | < 0.001 |
| Image Questions | 31/40 (77.5%) | 17/40 (42.5%) | 0.003 |
| **Subspeciality** |  |  |  |
| CKD/ESKD | 34/45 (75.6%) | 19/45 (42.2%) | 0.003 |
| AKI | 9/10 (90.0%) | 4/10 (40.0%) | 0.061 |
| Glomerular Diseases | 47/57 (82.5%) | 29/57 (50.9%) | < 0.001 |
| Tubulointerstitial Diseases | 17/19 (89.5%) | 9/19 (47.4%) | 0.015 |
| Hypertension/Vascular Diseases | 13/18 (72.2%) | 8/18 (44.4%) | 0.176 |
| Water/Electrolytes/<br>Acid-Base Disorder | 17/23 (73.9%) | 17/23 (73.9%) | 1.000 |
| ADPKD/Urology | 10/12 (83.3%) | 7/12 (58.3%) | 0.369 |
| Basic Medicine | 23/25 (92.0%) | 14/25 (56.0%) | 0.010 |

The performance of o1 pro and GPT-4 is reported for each category of Self-Assessment
Questions for Nephrology Board Renewal. Differences in performance between large language models were queried using chi-squared and Fisher's exact tests. CKD, chronic kidney disease; ESKD, end-stage kidney disease; AKI, acute kidney injury; ADPKD, autosomal dominant polycystic kidney disease.

Regarding question type, o1 pro also outpaced GPT-4. For general questions, o1 pro’s correct rate was 83.8% (93/111), exceeding GPT-4’s 49.5% (55/111; p<0.001). For clinical questions, o1 pro similarly excelled, with an accuracy of 78.6% (77/98) compared to GPT-4’s 53.1% (52/98; p<0.001).

Analysis by image inclusion showed that o1 pro demonstrated superior performance in both non-image and image-based questions. For non-image questions, o1 pro achieved 82.2% (139/169) and GPT-4 achieved 53.3% (90/169; p<0.001). For image-based questions, o1 pro’s correct rate was 77.5% (31/40) compared to GPT-4’s 42.5% (17/40; p=0.003).

Subspecialty analysis revealed that o1 pro significantly outperformed GPT-4 in several key areas, including CKD/ESKD (75.6% [34/45] vs. 42.2% [19/45]; p=0.003), glomerular diseases, tubulointerstitial diseases, and basic science. In the remaining domains (AKI, hypertension/vascular diseases, water/electrolyte/acid-base disorders, and ADPKD/urology), o1 pro’s accuracy equaled or exceeded that of GPT-4, although these differences did not reach statistical significance.

In sum, o1 pro consistently surpassed GPT-4 across virtually all examined domains— overall performance, annual pass rates, taxonomy levels, question types, presence or absence of images, and multiple nephrology subspecialties.

## DISCUSSION

This study showed that the new-generation LLM, o1 pro significantly outperformed GPT-4 on the comprehensive Japanese nephrology board renewal questions. Not only did o1 pro achieve passing scores in every examination year, but it also excelled in higher-order cognitive tasks such as clinical reasoning and interpretation of medical images. These results suggest that next-generation LLMs could extend beyond simple knowledge retrieval, integrating specialized medical knowledge, visual data processing, and context-specific decision-making into more advanced reasoning capabilities.

Previous studies have demonstrated the utility of LLMs in medical contexts, including GPT-4’s strong performance on general medical knowledge exams and national licensing examinations [12–15]. In nephrology, GPT-4 was able to pass most of the Polish nephrology specialty exams [19]. Additionally, our earlier study showed that GPT-4 significantly outperformed GPT-3.5 on the SAQ-NBR and met passing standards in several examination years [16]. However, this study is the first to demonstrate that o1 pro consistently surpasses GPT-4 on this rigorous, specialized assessment. Beyond knowledge accuracy, o1 pro maintained a robust performance in complex interpretive and clinical questions, suggesting a major improvement in integrating kidney disease pathophysiology. These advancements could result from expanded training data, architectural upgrades, reinforcement learning for factual consistency, and enhanced multimodal processing [20,21]. Further model updates may improve LLMs’ performance even more in nephrology.

A particularly notable finding was o1 pro’s superior performance on image-based questions, including radiological and pathological images. Interpreting such visual information requires both deep medical knowledge and clinical experience—points not often emphasized in prior LLM studies in nephrology [27]. Our results may indicate o1 pro’s potential as a multifaceted clinical tool in nephrology, where expertise with textual, numerical, and image-based data is essential. From an educational standpoint, these capabilities may enable more effective image-focused teaching and objective skill assessment. Based on previous studies demonstrating advancements in LLMs’ performance for analyzing radiological and pathological images [28–31], LLMs may have the potential to one day assist pathologists and nephrologists, improving the accuracy and consistency of medical imaging assessments.

These findings indicate that LLMs are moving closer to practical utility in nephrology education and clinical decision-making. Traditionally, clinicians have relied on textbooks, literature, lectures, and clinical training—yet the exponential growth of medical information makes it challenging to stay current [32]. LLMs like o1 pro may serve as on-demand knowledge resources, offering evidence summaries, restructured pathophysiological concepts, and assistance with image interpretation. A study showed that LLMs could improve clinicians’ exam performance in nephrology [33]. Given o1 pro’s high accuracy in both basic knowledge recall and complex problem-solving, it holds promise as a comprehensive educational and clinical support system for a broad range of users.

Despite o1 pro’s demonstrated advantages in numerous categories, its lack of performance improvement in the ‘water/electrolytes/acid-base disorder’ subspecialty is a notable exception. This is particularly striking given o1 pro’s design for enhanced inference, suggesting that the cognitive demands of these specific questions might differ. Often, electrolyte and acid-base problems rely on the application of well-established physiological principles, codified diagnostic algorithms, and readily accessible clinical guidelines [34–36]. Consequently, the type of complex, multi-step inference that o1 pro is optimized for might not be as critical in this domain, where GPT-4’s extensive knowledge base and recognition capabilities could be sufficient to achieve comparable accuracy, especially if the assessment emphasizes recall of diagnostic criteria or the application of standard treatment algorithms.

This study had several limitations. First, it focused on the Japan-specific nephrology questions, leaving questions about its applicability to other specialties, languages, or exam formats. Whether these results translate to clinical practice also remains unclear. Second, the internal reasoning processes of both o1 pro and GPT-4 were undisclosed [18,20], complicating efforts to pinpoint the exact mechanisms behind their performance differences. Third, the SAQ-NBR items were publicly available on the internet, so there is a possibility that o1 pro and GPT-4 might have encountered these questions during training. Given o1 pro’s knowledge cutoff in October 2023 and GPT-4’s in September 2021, data leakage may have occurred. To clarify, we have listed the publication dates of these questions and answers in Supplemental Table S2. Finally, variations in LLM outputs may arise from different prompts (“prompt engineering”), raising concerns about reproducibility. Furthermore, the inherent stochastic nature of LLMs, influenced by factors such as probabilistic sampling during response generation and the use of a temperature parameter to control randomness, can lead to different outputs even with the same input. This variability adds another layer of complexity when evaluating the performance and reliability of LLMs in specialized domains.

Future research should include cross-specialty and multimodal evaluations and large-scale analyses using varied exam formats, moving beyond multiple-choice questions to incorporate more clinically relevant formats such as free-response questions [37]. Comparisons with other models (e.g., Gemini, Claude, Llama) could clarify the relative advantages of different LLMs and contribute to the broader medical AI ecosystem.

Prospective clinical trials are needed to establish whether LLM integration in clinical workflows improves diagnostic and therapeutic outcomes and, most importantly, benefits patient care.

## CONCLUSION

This study suggested that o1 pro consistently surpassed GPT-4 in tackling diverse nephrology tasks, reflecting the increasingly sophisticated reasoning capabilities of next-generation LLMs. LLMs have the potential to support medical education and clinical decision-making in nephrology, leading to better patient care.

## ACKNOWLEDGEMENTS

None.

## AUTHORS’ CONTRIBUTIONS

R.N. designed the research plan. R.N., C.Y. and F.K. participated in collecting and analyzing the data about this study. R.N., D.I. and Y.S. participated in the writing of the paper. R.N., C.Y., F.K., D.I., Y.S. participated in the approval of the final manuscript.

## DATA AVAILABILITY

Due to the proprietary nature of the data used for this study (Self-Assessment Questions for Nephrology Board Renewal), the authors cannot post the raw data used for the analysis. However, the authors are able to share a part of the collected data (ex. large language model responses, etc.) on request to other researchers who have access to this exam. For data requests, please contact the corresponding author, Ryunosuke Noda.

## ETHICS APPROVAL AND CONSENT TO PARTICIPATE

This study did not involve human participants, human tissue samples, or any experiments on humans. Therefore, informed consent was not applicable. We consulted with the Division of Graduate Student Affairs and Research Promotion, which serves as the Institutional Review Board (IRB) office at St. Marianna University Hospital. After careful review, it was concluded that IRB approval was not indicated or required for this study because the study did not involve human subjects and is outside the scope of research requiring ethical review.

## CONSENT FOR PUBLICATION

Not applicable.

